# Protocol for the development and validation of a Rheumatoid Arthritis PredIction moDel using primary care health records (RAPID)

**DOI:** 10.1101/2024.04.09.24305328

**Authors:** Ben Hammond, Aliaksandra Baranskaya, Nicola Adderley, Dawit Zemedikun, Alexander d’Elia, Marie Falahee, Christian Mallen, Elspeth Insch, Joht Singh Chandan, Krishnarajah Nirantharakumar, Kym Snell, Karim Raza

## Abstract

**Background:** Rheumatoid Arthritis (RA) is a chronic rheumatological condition which causes inflammation of both the joint lining and extra-articular sites. It affects around 1% of the UK population and, if not properly treated, can lead joint damage, disability, and significant socioeconomic burden. The risk of long-term damage is reduced if treatment is started in an early disease stage with treatment in the first 3 months being associated with significantly improved clinical outcomes. However, treatment is often delayed due to long referral waits and challenges in identifying early RA in primary care. We plan to use large primary care datasets to develop and validate an RA risk prediction model for use in primary care, with the aim to provide an additional mechanism for early diagnosis and referral for treatment.

**Methods:** We identified candidate predictors from literature review, expert clinical opinion, and patient research partner input. Using coded primary care data held in Clinical Practice Research Datalink (CPRD) Aurum, we will use a time to event Cox proportional hazards model to develop a 1-year risk prediction model for RA. This will be validated first in CPRD GOLD and then independently in the Secure Anonymised Information Linkage dataset. We will also conduct a sensitivity analysis for the same model at 2–5-year risk, with a secondary outcome of RA and initiation of a disease modifying drug, and with the addition of laboratory test results as candidate predictors.

**Discussion:** The resulting risk prediction model may provide an additional mechanism to distinguish early RA in primary care and reduce treatment delays through earlier referral.

## Introduction

Rheumatoid Arthritis (RA) is a chronic rheumatological condition associated with both inflammation of the joint lining as well as wider extra-articular inflammation. (1,2) It currently affects approximately 1% of the UK population (3) and can present with a variety of symptoms including joint pain, stiffness swelling and fatigue. (4) If it is not treated effectively, it can lead to long term complications such as joint damage and disability, contribute to additional co-morbidities such as cardiovascular disease, and create a significant socioeconomic burden due to factors such as patients being unable to work. (5) However, very early treatment of rheumatoid arthritis (RA) has been shown to bring significant benefits, with treatment in the first 3 months of the condition being especially important. (6,7) Prior research has established that treatment within this period can lead to significantly improved clinical outcomes such as reduced joint damage, lower radiographic progression, and the potential for increased likelihood of sustained remission. (6,7)

Despite this, identifying patients in this early disease stage is challenging and many do not receive treatment within the recommended timeframe. (8) Musculoskeletal problems, such as back pain and osteoarthritis, present commonly in primary care (9) and distinguishing early RA symptoms from other musculoskeletal symptoms can be challenging for non-specialists. (10,11) Delays to early treatment are further confounded by long referral waiting times and a lack of fast-track pathways, with the 2021-2022 National Early Inflammatory Arthritis Audit finding that only 42% of patients received specialist review within 3 weeks of referral. (12)

Prediction models have been developed to predict adverse outcomes, treatment, and clinical response in RA patients, (13–19) however, to the best of our knowledge, no prediction models have yet utilised large primary care datasets to assess RA risk in a primary care setting. Such a prediction model could aid early diagnosis by flagging patients who are at a higher risk of having the condition as well as providing an additional mechanism to distinguish possible RA patients from those with similarly presenting common musculoskeletal pathologies.

Earlier work has established the presence of signs and symptoms associated with RA coded in large primary care database of electronic healthcare records, specifically Clinical Practice Research Datalink (CPRD). (20) Thus, we hypothesise that a prediction model trained on large, representative, primary care EHR datasets could provide better detection of early RA in primary care, better decision support for early referral to rheumatology specialist care and reduced treatment delay.

## Objectives

i. Develop and internally validate a prediction model for new diagnosis of RA in CPRD Aurum:

a. Using coded clinical diagnosis, symptoms, medications, and baseline patient characteristics as candidate predictors
b. Additionally using laboratory test results
ii. Externally validate the resulting prediction models in the CPRD GOLD and SAIL databases.
iii. Compare the performance of the models internally and externally using measures of calibration, discrimination, and clinical utility.

## Research Design and Methods

### Data sources

Routinely collected primary care data from three large anonymised electronic healthcare record (EHR) databases will be utilised. The first, CPRD Aurum, will be used to develop the models. CPRD Gold will then be used for initial external validation and SAIL will be used for independent external validation. These databases are described in further detail below:

1. Clinical Practice Research Datalink (CPRD) is an anonymised store of primary care records available for use in research. (21) It contains data on diagnosis, symptoms, medication, and laboratory test results as well as wider sociodemographic descriptors and baseline characteristics. (21) It is split into two databases, CPRD Aurum and Gold. CPRD Aurum includes practices using the EMIS system and primarily consists of practices in England and Northern Ireland and contains 16,011,762 active patients and 19.77% of UK general practices (as of December 2023). (22) CPRD GOLD includes those using the Vision software and mostly contains practices in Scotland. This provides data from an additional 2,967,792 patients covering another 4.55% of general practices. (23)
2. Secure Anonymised Information Linkage (SAIL) is an anonymised, Wales wide research available dataset containing EHR data representing 80% of the Welsh population, containing records of over 5 million patients who have used public services in Wales. (24–26)

### Target Population

This study will take a population level approach, specifically focussing on primary care. Patients are eligible for inclusion if they are aged 18 or over at index date (defined below), do not have a record of RA prior to index date and were registered with a GP practice contributing to the relevant database between 1^st^ January 2000 and 31^st^ December 2022. Additionally, patients must be registered at the practice for at least 12 months before being eligible to contribute data.

The aim of the prediction models is to aid referral and diagnosis of RA earlier in the disease pathway and so it is intended for use with individuals consulting with early symptoms of RA. Therefore, entry into the cohort will require patients to report at least one musculoskeletal trigger symptom, which are outlined in further detail below.

The index date will be the point a patient reports one of the trigger symptoms. Additional clinical codes reported 3 months after index will also be included as baseline values as it is likely there will be a delay for some additional symptoms to be recorded in the routine dataset. Follow up will be from the defined index date to the earliest of date of outcome, date of transfer to another practice, practice stops contributing to the dataset, study end date or death date.

### Study Outcome

The study outcome is defined as the earliest recorded diagnosis of rheumatoid arthritis occurring after the index date. This will be detected using a combination of SNOMED and Read codes (clinical coding systems used in primary care electronic healthcare systems in the NHS (27)) with the primary outcome being presence of a clinical code in the patient record indicating RA. A secondary outcome of a clinical code of RA as well as initiation of a disease-modifying antirheumatic drug (DMARD) will also be investigated. The lists of relevant codes defining the outcome will be reviewed by clinical members of the research team to ensure they accurately represent the outcome of interest.

### Clinical Predictor Variables

Initial work has selected candidate predictors through a multi-stage process. In the first step, a longlist of predictors of RA were identified from a literature review, clinical expert opinion from both General Practitioners and Rheumatologists, and patient research partner input. Predictors will be identified from codes contained in the patient record data so lists of SNOMED and Read codes have been created for each included predictor within the longlist. This resulted in a high number of predictors which could have reduced model stability and useability of any subsequent web calculator tools.

As a result, a second review was undertaken to reduce predictor numbers whilst retaining the most relevant clinical factors. To inform this, descriptive statistics were initially reported for each of the longlisted predictors. Non-musculoskeletal symptom codes with a clear clinical grouping will be first grouped into one predictor. Predictors with very low frequencies will then be grouped with similar codes if they share a clinically plausible link. These groups will be reviewed by clinician experts and any disagreements will be resolved by KR, a subject expert.

The candidate predictors which have been selected for use in the model development are reported in table 2. To ensure musculoskeletal and acute symptoms are related to a possible RA diagnosis, some predictor values are limited to only those reported up to 2 years prior to the index date. These were selected by clinical consensus and are also outlined in table 2. For patient characteristics and chronic conditions, no time limit was applied.

A subset of these predictors, reported in table 1 below, will additionally be used to define entry into the cohort. Predictors are eligible to be selected for this if they had a strong clinical association with RA, with a specific focus on selecting cardinal musculoskeletal symptoms of the condition. The initial list of predictors was presented to three clinicians, covering rheumatology and primary care, who independently selected the clinical symptoms they believed to have strong associations with RA that should trigger the model. Any conflicting results are resolved through a discussion with the study team, including KR who is a subject expert in RA. This process resulted in the trigger symptoms shown in Table 1, being selected as an additional entry criterion to the cohort.

**Table 1:** Symptoms that define entry into the cohort.

| Symptom | Description |
| --- | --- |
| Synovitis | A reported symptom of synovitis. |
| Arthralgia | A reported symptom of arthralgia or related synonyms. |
| Stiffness | A reported symptom of generalised stiffness or joint specific stiffness. Grouped due to low frequency of joint specific stiffness. |
| Swelling | A reported symptom of generalised swelling or joint specific swelling. Grouped due to low frequency of joint specific stiffness. |
| Palindromic RA | A record relating to palindromic RA. |
| Tendinitis | A reported symptom of tendinitis |
| Carpal Tunnel Syndrome | A reported symptom of carpal tunnel syndrome |
| Ankle Symptom (other or not specified) | Ankle related pathologies not listed below, or ankle mentioned but specific condition not otherwise specified. |
| Ankle Pain | A reported symptom of pain specifically linked to the ankle. |
| Ankle Arthritis | A reported symptom of a form of ankle arthritis not exclusive to rheumatoid arthritis. |
| Elbow Symptom (other or not specified) | Elbow related pathologies not listed below, or elbow mentioned but specific condition not otherwise specified. |
| Elbow Pain | A reported symptom of pain specifically linked to the elbow. |
| Elbow Arthritis | A reported symptom of a form of elbow arthritis not exclusive to rheumatoid arthritis. |
| Foot Symptom (other or not specified) | Foot related pathologies not listed below, or foot mentioned but specific condition not otherwise specified. |
| Foot Pain | A reported symptom of pain specifically linked to the foot. |
| Foot Arthritis | A reported symptom of a form of foot arthritis not exclusive to rheumatoid arthritis. |
| Jaw Symptom (other or not specified) | Jaw related pathologies not listed below, or jaw mentioned but specific condition not otherwise specified. |
| Jaw Pain | A reported symptom of pain specifically linked to the jaw. |
| Knee Symptom (other or not specified) | Knee related pathologies not listed below, or knee mentioned but specific condition not otherwise specified. |
| Knee Pain | A reported symptom of pain specifically linked to the knee. |
| Knee Arthritis | A reported symptom of a form of knee arthritis not exclusive to rheumatoid arthritis. |
| Shoulder Symptom (other or not specified) | Shoulder related pathologies not listed below, or shoulder mentioned but specific condition not otherwise specified. |
| Shoulder Pain | A reported symptom of pain specifically linked to the shoulder. |
| Shoulder Arthritis | A reported symptom of a form of shoulder arthritis not exclusive to rheumatoid arthritis. |
| Wrist Symptom (other or not specified) | Wrist related pathologies not listed below, or wrist mentioned but specific condition not otherwise specified. |
| Wrist Pain | A reported symptom of pain specifically linked to the wrist. |
| Wrist Arthritis | A reported symptom of a form of wrist arthritis not exclusive to rheumatoid arthritis. |
| Neck Other | Neck related pathologies not listed below, or neck mentioned but specific condition not otherwise specified. |
| Neck Pain | A reported symptom of pain specifically linked to the neck. |
| Hand Other | Hand related pathologies not listed below, or hand mentioned but specific condition not otherwise specified. |
| Hand Pain | A reported symptom of pain specifically linked to the hand. |
| Hand Arthritis | A reported symptom of a form of hand arthritis not exclusive to rheumatoid arthritis. |
| Hip Other | Hip related pathologies not listed below, or hip mentioned but specific condition not otherwise specified. |
| Hip Pain | A reported symptom of pain specifically linked to the hip. |
| Hip Arthritis | A reported symptom of a form of hip arthritis not exclusive to rheumatoid arthritis. |

**Table 2:** List of predictors and their period of eligibility for inclusion.

| Final Code List Name | Longlist Code List Name | Description | Timeline |
| --- | --- | --- | --- |
| **Musculoskeletal Symptom or Diagnosis** |  |  |  |
| Muscle Pain and Cramps | Muscle Pain and Cramps | Record of either muscle pain or cramps | 2-Years |
| Stiffness | Stiffness | Record of generalised stiffness or joint specific stiffness. Grouped due to low frequency of joint specific stiffness. | 2-Years |
|  | Ankle Stiffness |  |  |
|  | Elbow Stiffness |  |  |
|  | Foot Stiffness |  |  |
|  | Jaw Stiffness |  |  |
|  | Knee Stiffness |  |  |
|  | Shoulder Stiffness |  |  |
|  | Wrist Stiffness |  |  |
|  | Neck Stiffness |  |  |
|  | Hand Stiffness |  |  |
|  | Hip Stiffness |  |  |
| Swelling | Ankle Swelling | Record of generalised swelling or joint specific swelling. Grouped due to low frequency of joint specific stiffness. | 2-Years |
|  | Elbow Swelling |  |  |
|  | Foot Swelling |  |  |
|  | Jaw Inflammation |  |  |
|  | Knee Swelling |  |  |
|  | Shoulder Swelling |  |  |
|  | Wrist Swelling |  |  |
|  | Hand Swelling |  |  |
|  | Hip Swelling |  |  |
| Tendinitis | Tendinitis | Record of tendinitis | 2-Years |
| Synovitis | Synovitis | Record of synovitis | 2-Years |
| Altered Sensation | Altered Sensation | Record of altered sensation or related synonyms. | 2-Years |
| Arthralgia | Arthralgia | Record of arthralgia or related synonyms. | 2-Years |
| Carpal Tunnel Syndrome | Capel Tunnel Syndrome | Record of carpal tunnel syndrome | 2-Years |
| Family History of RA | Family History of RA | Record of family history of RA. | Lifetime |
| Ankle Other | Ankle Abnormal Finding but Not Otherwise Specified | Ankle related pathologies not listed below, or ankle mentioned but specific condition not otherwise specified. | 2-Years |
|  | Ankle Enthesopathy |  |  |
|  | Ankle Ganglion |  |  |
|  | Ankle Impingement |  |  |
|  | Ankle Sprain |  |  |
|  | Ankle Tendinitis |  |  |
|  | Ankle Normal Finding but Not Otherwise Specified |  |  |
|  | Ankle Mentioned but Not Otherwise Specified (Neutral) |  |  |
| Ankle Pain | Ankle Pain | Record of pain specifically linked to the ankle. | 2-Years |
| Ankle Arthritis | Ankle Arthritis | Record of a form of ankle arthritis not exclusive to rheumatoid arthritis. | 2-Years |
| Elbow Other | Elbow Ganglion | Elbow related pathologies not listed below, or elbow mentioned but specific condition not otherwise specified. | 2-Years |
|  | Elbow Abnormal Finding but Not Otherwise Specified |  |  |
|  | Elbow Bursitis |  |  |
|  | Elbow Cubital Tunnel Syndrome |  |  |
|  | Elbow Enthesopathy |  |  |
|  | Elbow Epicondylitis |  |  |
|  | Elbow Sprain |  |  |
|  | Elbow Mentioned but Not Otherwise Specified (Neutral) |  |  |
|  | Elbow Normal Finding but Not Otherwise Specified |  |  |
| Elbow Pain | Elbow Pain | Record of pain specifically linked to the elbow. | 2-Years |
| Elbow Arthritis | Elbow Arthritis | Record of a form of elbow arthritis not exclusive to rheumatoid arthritis. | 2-Years |
| Foot Other | Foot Bursitis | Foot related pathologies not listed below, or foot mentioned but specific condition not otherwise specified. | 2-Years |
|  | Foot Enthesopathy |  |  |
|  | Foot Fasciitis |  |  |
|  | Foot Ganglion |  |  |
|  | Foot Morton's |  |  |
|  | Foot Sprain |  |  |
|  | Foot Tendinitis |  |  |
|  | Foot Tarsal Tunnel Syndrome |  |  |
|  | Foot Mentioned but Not Otherwise Specified (Neutral) |  |  |
|  | Foot Normal Finding but Not Otherwise Specified |  |  |
|  | Foot Abnormal Finding but Not Otherwise Specified |  |  |
| Foot Pain | Foot Pain | Record of pain specifically linked to the foot. | 2-Years |
| Foot Arthritis | Foot Arthritis | Record of a form of foot arthritis not exclusive to rheumatoid arthritis. | 2-Years |
| Jaw Other | Jaw Abnormal Finding but Not Otherwise Specified | Jaw related pathologies not listed below, or jaw mentioned but specific condition not otherwise specified. | 2-Years |
|  | Jaw Disorder |  |  |
|  | Jaw Temporomandibular Joint Dysfunction |  |  |
|  | Jaw Mentioned but Not Otherwise Specified (Neutral) |  |  |
|  | Jaw Normal Finding but Not Otherwise Specified |  |  |
| Jaw Pain | Jaw Pain | Record of pain specifically linked to the jaw. | 2-Years |
| Knee Other | Knee Abnormal Finding but Not Otherwise Specified | Knee related pathologies not listed below, or knee mentioned but specific condition not otherwise specified. | 2-Years |
|  | Knee Bursitis |  |  |
|  | Knee Enthesopathy |  |  |
|  | Knee Iliofemoral |  |  |
|  | Knee Patellofemoral Pathology |  |  |
|  | Knee Sprains |  |  |
|  | Knee Tenosynovitis |  |  |
|  | Knee Mentioned but Not Otherwise Specified (Neutral) |  |  |
|  | Knee Normal Finding but Not Otherwise Specified |  |  |
| Knee Pain | Knee Pain | Record of pain specifically linked to the knee. | 2-Years |
| Knee Arthritis | Knee Arthritis | Record of a form of knee arthritis not exclusive to rheumatoid arthritis. | 2-Years |
| Shoulder Other | Shoulder Ganglion | Shoulder related pathologies not listed below, or shoulder mentioned but specific condition not otherwise specified. | 2-Years |
|  | Shoulder Abnormal Finding but Not Otherwise Specified |  |  |
|  | Shoulder Bursitis |  |  |
|  | Shoulder Frozen |  |  |
|  | Shoulder Impingement |  |  |
|  | Shoulder Abnormal Finding but Not Otherwise Specified |  |  |
|  | Shoulder Sprain |  |  |
|  | Shoulder Tenosynovitis |  |  |
|  | Shoulder Mentioned but Not Otherwise Specified (Neutral) |  |  |
|  | Shoulder Normal Finding but Not Otherwise Specified |  |  |
| Shoulder Pain | Shoulder Pain | Record of pain specifically linked to the shoulder. | 2-Years |
| Shoulder Arthritis | Shoulder Arthritis | Record of a form of shoulder arthritis not exclusive to rheumatoid arthritis. | 2-Years |
| Wrist Other | Wrist Abnormal Finding but Not Otherwise Specified | Wrist related pathologies not listed below, or wrist mentioned but specific condition not otherwise specified. | 2-Years |
|  | Wrist Bursitis |  |  |
|  | De Quervain's Tenosynovitis |  |  |
|  | Wrist Enthesopathy |  |  |
|  | Wrist Sprains |  |  |
|  | Wrist Tenosynovitis |  |  |
|  | Wrist Ganglion |  |  |
|  | Wrist Mentioned but Not Otherwise Specified (Neutral) |  |  |
|  | Wrist Normal Finding but Not Otherwise Specified |  |  |
| Wrist Pain | Wrist Pain | Record of pain specifically linked to the wrist. | 2-Years |
| Wrist Arthritis | Wrist Arthritis | Record of a form of wrist arthritis not exclusive to rheumatoid arthritis. | 2-Years |
| Neck Other | Neck Abnormal Finding but Not Otherwise Specified | Neck related pathologies not listed below, or neck mentioned but specific condition not otherwise specified. | 2-Years |
|  | Neck Mentioned but Not Otherwise Specified (Neutral) |  |  |
|  | Neck Pathology Reported |  |  |
|  | Neck Sprain |  |  |
|  | Neck Normal Finding but Not Otherwise Specified |  |  |
| Neck Pain | Neck Pain | Record of pain specifically linked to the neck. | 2-Years |
| Hand Other | Hand Ganglion | Hand related pathologies not listed below, or hand mentioned but specific condition not otherwise specified. | 2-Years |
|  | Hand Abnormal Finding but Not Otherwise Specified |  |  |
|  | Hand Bursitis |  |  |
|  | Hand Mentioned but Not Otherwise Specified |  |  |
|  | Hand Sprain |  |  |
|  | Hand Tendinitis |  |  |
|  | Make A Fist |  |  |
|  | Hand Mentioned but Not Otherwise Specified (Neutral) |  |  |
|  | Hand Normal Finding but Not Otherwise Specified |  |  |
| Hand Pain | Hand Pain | Record of pain specifically linked to the hand. | 2-Years |
| Hand Arthritis | Hand Arthritis | Record of a form of hand arthritis not exclusive to rheumatoid arthritis. | 2-Years |
| Hip Other | Hip Ganglion | Hip related pathologies not listed below, or hip mentioned but specific condition not otherwise specified. | 2-Years |
|  | Hip Abnormal Finding but Not Otherwise Specified |  |  |
|  | Hip Bursitis |  |  |
|  | Hip Enthesopathy |  |  |
|  | Hip Impingement |  |  |
|  | Hip Irritable |  |  |
|  | Hip Sprain |  |  |
|  | Hip Mentioned but Not Otherwise Specified (Neutral) |  |  |
|  | Hip Normal Finding but Not Otherwise Specified |  |  |
| Hip Pain | Hip Pain | Record of pain specifically linked to the hip. | 2-Years |
| Hip Arthritis | Hip Arthritis | Record of a form of hip arthritis not exclusive to rheumatoid arthritis. | 2-Years |
| **Non-Musculoskeletal Clinical Diagnosis** |  |  |  |
| Viral Infection | Record of EBV Infection | Infection with EBV or parvovirus | 2-Years |
|  | Record of Parvovirus Infection |  |  |
| Heart Disease | Record of IHD | Record of an MI or heart failure | Lifetime |
|  | Record of an MI |  |  |
|  | Record of Heart Failure |  |  |
| Stroke | Ischaemic Stroke | Record of stroke | Lifetime |
|  | Haemorrhagic Stroke |  |  |
|  | Unspecified Stroke |  |  |
| Allergies | Allergies | Record of allergies | Lifetime |
| Atopic Conditions | Asthma | Record of atopic asthma, eczema, or allergic rhinitis | Lifetime |
|  | Atopic Eczema |  |  |
|  | Allergic Rhinitis |  |  |
| Mental Health Condition | Depression | Record of anxiety, depression, or PTSD | Lifetime |
|  | Anxiety |  |  |
|  | PTSD |  |  |
| IBD | Crohn's Disease | Record of Crohn's or Ulcerative Colitis | Lifetime |
|  | Ulcerative Colitis |  |  |
| Embolism | Pulmonary Embolism | Record of either venous or pulmonary thromboembolism | Lifetime |
|  | Venous thromboembolism |  |  |
| Lung Pathology | Interstitial Lung Disease | Record of COPD or interstitial lung disease | Lifetime |
|  | COPD |  |  |
| Type 1 DM | Type 1 Diabetes Mellitus | Record of Type 1 diabetes mellites | Lifetime |
| Type 2 DM | Type 2 Diabetes Mellitus | Record of Type 1 diabetes mellites | Lifetime |
| Autoimmune Thyroid | Autoimmune Thyroid | Record of autoimmune thyroid disease | Lifetime |
| Endometriosis | Endometriosis | Record of endometriosis | Lifetime |
| Early Menopause | Early Menopause | Record of early menopause | Lifetime |
| Hepatitis | Hepatitis C | Record of hepatitis | Lifetime |
| Pemphigus | Pemphigus | Record of pemphigus | lifetime |
| Periodontitis | Peritonitis | Record of periodontitis | Lifetime |
| Pregnancy | Pregnancy | Record of either pregnancy or pregnancy exclusive condition/ symptom. | Lifetime |
| Sleep Problems | Sleep Problems | Record suggesting insomnia or related synonyms | 2-Years |
| Stress | Stress | Record suggesting high stress levels | Lifetime |
| **Other Clinical Findings** |  |  |  |
| Weakness | Weakness | Record of weakness or related synonyms. | 2-Years |
| Falls | Falls | Record of falls | 2-Years |
| Fatigue | Fatigue | Record of fatigue, tiredness, or related synonyms | 2-Years |
| Weight Loss | Weight loss | Record of unexpected weight loss. | 2-Years |
| **Medication Use** |  |  |  |
| NSAID Use | Topical NSAID Use | Use of NSAID | 2-Years |
|  | Topical NSAID Derivatives |  |  |
|  | Oral NSAID |  |  |
| Opioid Use | Weak Opioid Use | Use of opioid | 2-Years |
|  | Strong Opioid Use |  |  |
| Oral Corticosteroid Use | Systemic Oral corticosteroid Use | Use of oral corticosteroids | 2-Years |
| Contraceptives | Progesterone Only Contraceptive Pill use | Use of either combined or progesterone only pill | Lifetime |
|  | Combined oral contraceptive pill use |  |  |
| Anti-Depressant Use | SSRI Use | Use of an antidepressant | 2-Years |
|  | Amitriptyline Use |  |  |
|  | SNRI use |  |  |
| Hormone Replacement Therapy | Hormone Replacement Therapy | Use of hormone replacement therapy | Lifetime |
| Statins | Statins | Use of statins | Lifetime |
| Vitamin D use | Vitamin D use | Use of vitamin D | Lifetime |
| **Laboratory Tests** |  |  |  |
| Haemoglobin | Haemoglobin | Serum haemoglobin count | 2-Years |
| Platelet Count | Platelet Count | Serum platelet count | 2-Years |
| Erythrocyte Sedimentation Rate | Erythrocyte Sedimentation Rate | Erythrocyte Sedimentation Rate | 2-Years |
| C-Reactive Protein | C-Reactive Protein | Serum CRP level | 2-Years |
| HbA1C | HbA1C | Serum HbA1C level | 2-Years |
| Rheumatoid Factor | Rheumatoid Factor | Serum Rheumatoid Factor Level | 2-Years |
| eGFR | eGFR | eGFR rate recorded in record | 2-Years |
| Vitamin D Level | Vitamin D Level | Serum vitamin D level | 2-Years |
| Anti-Cyclic Citrullinated Peptide | Anti-Cyclic Citrullinated Peptide | Serum anti-CCP level | 2-Years |
| **Baseline Characteristics** |  |  |  |
| Age | Age | Participant age at baseline | Lifetime |
| Sex | Sex | Patient sex | Lifetime |
| Ethnicity | Ethnicity | Patient ethnicity (White, Black, Asian, Other and Mixed, Missing) | Lifetime |
| BMI | BMI | Patient BMI as continuous value | Lifetime |
| Index of Multiple Deprivation Score | Index of Multiple Deprivation Score | Participant IMD score at postcode level | Lifetime |
| Smoking Status | Smoking Status | Current patient tobacco smoking status (Smoker, Ex-Smoker, Non-Smoker) | Lifetime |
| Alcohol Misuse | Alcohol Misuse | Record of alcohol misuse or excess alcohol use. | Lifetime |
| High levels of Physical Activity | High levels of physical activity | Record reporting high levels of physical activity or exercise. | Lifetime |
| Low levels of activity | Low levels of physical activity | Record reporting low levels of physical activity or exercise | Lifetime |

## Statistical analysis

### Descriptive Statistics

To summarise the cohorts for both the model training and validations, we will report descriptive statistics. Categorical and binary variables will be summarised using frequencies and percentages. Continuous variables will be summarised by mean and standard deviation when normally distributed or median and interquartile range when not. A baseline table describing the cohort characteristics, including stratifying by those who did and did not develop the outcome of interest within the study period, will also be reported.

### Missing data

The degree of missing data for each candidate predictor will be investigated prior to model development. For each predictor, descriptive statistics will be reported as described above. Several separate approaches will then be utilised to handle missing data depending on missingness and clinical importance. In a similar approach as reported in other EHR based prediction models (28), the absence of data relating to a clinical condition or symptom recorded in a binary format will be presumed to mean that the condition is not present for that participant. For categorical predictors (e.g. smoking status and ethnicity) a separate missing category will be created. For continuous predictors, suitable imputation strategies will be investigated as, due to the nature of EHR, missingness is likely informative for these variables and this will need to be considered within any imputation approach.

### Model Development

Predictor selection will be carried out using multivariable fractional polynomial models. As part of this, we will carry out backwards elimination with predictors not meeting the 0.157 level of significance being removed from the model. Continuous variables will be kept as continuous and modelled non-linearly using fractional polynomials when the fit is improved. Clinically significant variables, decided by clinical expert opinion, will be forced into the model regardless of statistical significance. All models will utilise Cox Proportional Hazards Regression. The proportional hazards assumptions will be checked using ‘log-log’ plots and an extension to time dependent effects will be considered if necessary.

### Internal Validation

Due to the large dataset, overfitting and optimism are expected to be very small (see sample size section below). Bootstrapping would be computationally intensive and only provide small adjustments. As such, a heuristic uniform shrinkage factor will be calculated and used to adjust for any optimism present in the final model if necessary.

### Model Performance

Model fit will be appraised using Cox-Snell and Nagelkerke R-squared. Model performance will be evaluated using measures of discrimination and calibration. Discrimination will be evaluated using Harrell’s C index and assessed over the whole follow-up period as well as looking at time-dependent C indexes. Calibration will be evaluated by plotting predicted and observed probability of the outcome (calibration plot), ratio of observed and expected outcome (calibration in the large) and the calibration slope. Clinical utility will be assessed using decision curve analysis, showing the net benefit of the model across a range of threshold probabilities. All performance measures will be calculated at the primary point of follow up of 1 year.

### Sensitivity Analysis

Model performance will be analysed using both the primary outcome definition of RA, defined by an RA code, as well as by a sensitivity analysis including the secondary outcome of RA defined by both an RA code and evidence of starting a DMARD. Model performance will also be evaluated at 2–5-year risk through adjusting the baseline survival value, in addition to the primary follow up of 1 year.

### Sample Size for Development

Utilising an estimated event rate of 0.0017, a target shrinkage factor of 0.9, 95 predictor parameters, and an R^2^ value of 0.15 (default), we estimated that, for predictions at 1 year after the index date, the minimum sample size required would be 213,275 patients with 363 outcome events. Due to CPRD Aurum containing 16,011,762 active patients (22), we believe we will very likely exceed this requirement and anticipate low overfitting. If this is not the case, we will reduce the number of model parameters to ensure the expected shrinkage is no less than 0.9.

### External Validation

External validation of the final model will be carried out in two separate datasets. First, the model will be externally validated in CPRD GOLD by the University of Birmingham research group who will also be developing the model. Secondly, an independent group at the University of Swansea will validate the model in the SAIL dataset. Due to size of the training and validation datasets, we will use the same methods for internal and external validation as we don’t expect a large degree of optimism.

### Sample Size for External Validation

Sample size calculations for external validation have not been formally calculated as, given the expected size of cohort, a sufficient amount of data should be available for precise estimates of model performance.

### Statistical Software

Analysis and data preparation will be caried out using the software packages Python (v3.10.8) and Stata (v18).

### Model Presentation

The resulting model equations will be reported in the resulting manuscript and will also be presented as an interactive web calculator to aid with dissemination and implementation of the model.

## Discussion

This study aims to develop and validate a risk prediction model for RA in primary care. It will provide an opportunity to utilise large primary care datasets for both model development and validation as such can provide several benefits. The training data should enable a model to be generalisable to the intended use case of point of care in primary care. The proposed datasets should translate into a large sample size and thus allow for more stable model parameters. (29) The model will also be externally validated in two separate databases, with one being carried out by an independent group. This should provide a high level of validation and establish increased generalisability to UK primary care.

This study will, however, have limitations. Early analysis has suggested the possibility of poor coding of symptoms. Joint specific pathology appears to be poorly reported and may be more prevalent in free text notes, which we cannot access in the present study. Future work using natural language processing may provide a mechanism to address this and further increase the accuracy of future models. (30) Furthermore, we envisage missing data to be a significant issue, especially for continuous variables. However, we will have missing data strategies which aim to reduce the effects of this where possible. Finally, although the dataset is representative of primary care, coding practices can vary between GP practices and as such local performance of the model could vary. (31) Additionally, if a model was adopted this may further change coding practices and future retraining of a model may be required.

## Data Availability

Data access will be subject to ethics approval from CPRD.

## Declarations

### Ethics Approval

CPRD obtains annual research ethics approval from the UK’s Health Research Authority Research Ethics Committee (East Midlands, Derby; reference no.05/MRE04/87) to receive and supply patient data for research. Therefore, no additional ethics approval is required for studies using CPRD data for research, subject to individual research protocols meeting CPRD data governance requirements. The use of CPRD data for the study was approved by the CPRD Independent Scientific Advisory Committee (reference no. 22_002239). Individual patient data is available from CPRD with valid license.

### Patient Involvement

Development of this protocol as well as the ongoing prediction model is aided by monthly project meetings in which a patient partner, EI, participates and provides insight from a patient perspective.

### Funding

BH is funded by an MB-PhD studentship supported by The Kennedy Trust for Rheumatology Research [grant no. KENN 2021 04]. NIHR Research for Patient Benefit funds the Development and validation of Rheumatoid Arthritis PredIction moDel using primary care health records (RAPID), grant NIHR203621. AD is funded by a PhD studentship from the Applied Research Collaboration Northwest, in turn funded by the National Institute for Health Research (NIHR). KR, KN and NJA are supported by the NIHR Birmingham Biomedical Research Centre (BRC). This is independent research carried out at the NIHR BRC. The views expressed are those of the author(s) and not necessarily those of the NIHR or the Department of Health and Social Care. CM is part funded by the NIHR ARC West Midlands and the NIHR School for Primary Care Research

### Conflict of Interest

JSC and KN are co-directors of DExtER operating division which is part of the University of Birmingham. DExtER operating division supports the extraction and preparing of healthcare data to support epidemiological analyses such as those seen in this article.

## Notes

### Author Declarations

CPRD obtains annual research ethics approval from the UKs Health Research Authority Research Ethics Committee (East Midlands, Derby; reference no.05/MRE04/87) to receive and supply patient data for research. Therefore, no additional ethics approval is required for studies using CPRD data for research, subject to individual research protocols meeting CPRD data governance requirements. The use of CPRD data for the study was approved by the CPRD Independent Scientific Advisory Committee (reference no. 22_002239). Individual patient data is available from CPRD with valid license.

## References

1. Bullock J, Rizvi SAA, Saleh AM, Ahmed SS, Do DP, Ansari RA, et al. Rheumatoid Arthritis: A Brief Overview of the Treatment. Med Princ Pract. 2019; 27(6): 501–507

2. Smolen JS, Aletaha D, McInnes IB. Rheumatoid arthritis. The Lancet. 2016; 388(10055):2023–38.

3. Scott IC, Whittle R, Bailey J, Twohig H, Hider SL, Mallen CD, et al. Rheumatoid arthritis, psoriatic arthritis, and axial spondyloarthritis epidemiology in England from 2004 to 2020: An observational study using primary care electronic health record data. The Lancet Regional Health – Europe. 2022; 23.

4. Smolen JS, Aletaha D, McInnes IB. Rheumatoid arthritis. The Lancet. 2016; 388(10055):2023–38.

5. Wu D, Luo Y, Li T, Zhao X, Lv T, Fang G, et al. Systemic complications of rheumatoid arthritis: Focus on pathogenesis and treatment. Front Immunol. 2022; 13: 1051082

6. Raza K, Buckley CE, Salmon M, Buckley CD. Treating very early rheumatoid arthritis. Best Pract Res Clin Rheumatol. 2006; 20(5):849–63.

7. van der Linden MP, le Cessie S, Raza K, van der Woude D, Knevel R, Huizinga TW, et al. Long-term impact of delay in assessment of patients with early arthritis. Arthritis Rheum. 2010; 62(12):3537–46.

8. Chilton F, Bradley E, Mitchell T. ‘Lost time’. Patients with early inflammatory/rheumatoid arthritis and their experiences of delays in Primary Care. Musculoskeletal Care. 2021; 19(4):495–503.

9. Margham, T. Musculoskeletal disorders: time for joint action in primary care. British Journal of General Practice. 2011; 61(592): 657.

10. Stack RJ, van Tuyl LH, Sloots M, van de Stadt LA, Hoogland W, Maat B, et al. Symptom complexes in patients with seropositive arthralgia and in patients newly diagnosed with rheumatoid arthritis: a qualitative exploration of symptom development. Rheumatology (Oxford). 2014; 53(9):1646–53.

11. Suter LG, Fraenkel L, Holmboe ES. What factors account for referral delays for patients with suspected rheumatoid arthritis? Arthritis Rheum. 2006; 55(2):300–5.

12. Galloway J, Ledingham J, Coalwood C, Oyebanjo S, Garnavos N. National Early Inflammatory Arthritis Audit (NEIAA). [internet]. 2022 [cited 2024 Mar 10]. Available from: https://www.rheumatology.org.uk/Portals/0/Documents/Practice_Quality/Audit/NEIA/2022/NEIAA%20Fourth%20Annual%20Report_FINAL_11.01.23.pdf?ver=2023-01-11-165709-690

13. Lezcano-Valverde JM, Salazar F, León L, Toledano E, Jover JA, Fernandez-Gutierrez B, et al. Development and validation of a multivariate predictive model for rheumatoid arthritis mortality using a machine learning approach. Scientific Reports. 2017; 7(1):10189.

14. Yang C, Williams RD, Swerdel JN, Almeida JR, Brouwer ES, Burn E, et al. Development and external validation of prediction models for adverse health outcomes in rheumatoid arthritis: A multinational real-world cohort analysis. Semin Arthritis Rheum. 2022; 56:152050.

15. Archer R, Hock E, Hamilton J, Stevens J, Essat M, Poku E, et al. Assessing prognosis and prediction of treatment response in early rheumatoid arthritis: systematic reviews. Health Technol Assess. 2018; 22(66):1–294.

16. de Rotte M, Pluijm SMF, de Jong PHP, Bulatović Ćalasan M, Wulffraat NM, Weel A, et al. Development and validation of a prognostic multivariable model to predict insufficient clinical response to methotrexate in rheumatoid arthritis. PLoS One. 2018; 13(12):e0208534.

17. Scott IC, Seegobin SD, Steer S, Tan R, Forabosco P, Hinks A, et al. Predicting the Risk of Rheumatoid Arthritis and Its Age of Onset through Modelling Genetic Risk Variants with Smoking. PLOS Genetics. 2013; 9(9):e1003808.

18. van de Stadt LA, Witte Bi Fau - Bos WH, Bos Wh Fau - van Schaardenburg D, van Schaardenburg D. A prediction rule for the development of arthritis in seropositive arthralgia patients. Ann Rheum Dis. 2013; 72(12):1920–6

19. van der Helm-van Mil AH, Detert J, le Cessie S, Filer A, Bastian H, Burmester GR, et al. Validation of a prediction rule for disease outcome in patients with recent-onset undifferentiated arthritis: moving toward individualized treatment decision-making. Arthritis Rheum. 2008; 58(8):2241–7.

20. Muller S, Hider S, Machin A, Stack R, Hayward RA, Raza K, et al. Searching for a prodrome for rheumatoid arthritis in the primary care record: A case-control study in the clinical practice research datalink. Semin Arthritis Rheum. 2019; 48(5):815–20.

21. CPRD. CPRD Aurum Frequently asked questions (FAQs). [internet]. 2023 [cited 2024 Mar 10]. Available from: https://cprd.com/sites/default/files/2023-12/CPRD%20Aurum%20FAQs%20v2.4.pdf

22. CPRD. CPRD Aurum December 2023 Dataset. [internet]. 2023 [cited 2024 Mar 10]. Available from: https://cprd.com/cprd-aurum-december-2023-dataset

23. CPRD. CPRD GOLD December 2023 dataset [internet]. 2023 [cited 2024 Mar 10]. Available from: https://cprd.com/cprd-aurum-december-2023-dataset

24. Lyons J, Akbari A, Agrawal U, Harper G, Azcoaga-Lorenzo A, Bailey R, Rafferty J, et al. Protocol for the development of the Wales Multimorbidity e-Cohort (WMC): data sources and methods to construct a population-based research platform to investigate multimorbidity. BMJ Open. 2021; 11(1):e047101.

25. HDR UK. SAIL Databank [internet]. [date unknown] [cited 2024 Mar 10]. Available from: https://cprd.com/cprd-aurum-december-2023-dataset

26. Jones K. H, Ford D. V, Thompson S, Lyons R. A Profile of the SAIL Databank on the UK Secure Research Platform. International Journal of Population Data Science. 2020; 4(2).

27. CPRD. Defining your study population [internet]. 2023 [cited 2024 Mar 10]. Available from: https://cprd.com/defining-your-study-population

28. Wambua S, Crowe F, Thangaratinam S, O’Reilly D, McCowan C, Brophy S, et al. Protocol for development and validation of postpartum cardiovascular disease (CVD) risk prediction model incorporating reproductive and pregnancy-related candidate predictors. Diagnostic and Prognostic Research. 2022; 6(1):23.

29. Pate A, Emsley R, Sperrin M, Martin GP, van Staa T. Impact of sample size on the stability of risk scores from clinical prediction models: a case study in cardiovascular disease. Diagnostic and Prognostic Research. 2020; 4(1):14.

30. Ford E, Carroll J, Smith H, Daviers K, Koeling R, Petersen I, et al. What evidence is there for a delay in diagnostic coding of RA in UK general practice records? An observational study of free text. BMJ Open. 2016; 6(6):e010393.

31. Tai TW, Anandarajah S, Dhoul N, de Lusignan S. Variation in clinical coding lists in UK general practice: a barrier to consistent data entry? Inform Prim Care. 2007; 15(3):143–50.

